# Associations between perinatal biomarkers of maternal dairy fat intake and child cognitive development: results from the EDEN mother-child cohort

**DOI:** 10.1101/2024.05.03.24306802

**Authors:** Wen Lun Yuan, Martine Armand, Hugo Peyre, Catherine Sarté, Marie Aline Charles, Barbara Heude, Jonathan Y. Bernard

**Affiliations:** Université Paris Cité and Université Sorbonne Paris Nord, Inserm, INRAE, Center for Research in Epidemiology and StatisticS (CRESS), F-75004 Paris, France; Singapore Institute for Clinical Sciences, Agency for Science, Technology, and Research (A*STAR), Singapore, Singapore; Aix Marseille Univ, CNRS, CRMBM, Marseille, France; Centre de Ressources Autisme Languedoc-Roussillon et Centre d’Excellence sur l’Autisme et les Troubles Neuro-développementaux, CHU Montpellier, 34295 Montpellier cedex 05, France; Université Paris-Saclay, UVSQ, Inserm, CESP, Team DevPsy, 94807 Villejuif, France Team of Cognitive development and pathologies, Cognitive Sciences and Psycholinguistics Laboratory (LSCP), CNRS, 75005 Paris, France

## Abstract

**Background/Objective:** Offspring’s optimal neurodevelopment depends on maternal dietary lipids supply during the perinatal period. Dairy products are an important source of fat in Western diets. This study aimed at examining the associations between maternal dairy fat intake during pregnancy, evaluated through biomarkers in perinatal biofluids (C15:0 and C17:0) and child’s cognitive outcomes.

**Subjects/Methods:** Participants (N up to 1200) were French mother-child pairs from the EDEN cohort study. Biomarkers were assessed in the following biofluids: maternal red blood cells (RBC) membrane at 24 weeks’ gestation, cord RBC membrane and colostrum. Cognitive outcomes were language abilities scores at ages 2 and 3 years and overall cognitive development scores at ages 3 and 5-6 years. Associations were assessed using multiple linear regression models adjusted for factors related to family socioeconomic context, maternal lifestyle and healthy diet score. Interaction of any breastfeeding duration on the associations with dairy fat biomarkers in colostrum was studied.

**Results:** Positive associations were observed between levels of C17:0 in cord RBC and C15:0 in maternal RBC with language abilities. In children breastfed for at least 6 months, C15:0 level in colostrum was positively associated with language abilities at age 3 years and overall cognitive development at ages 3 and 5-6 years whereas C17:0 level was only associated with overall cognitive development at age 3 years.

**Conclusion:** Our results suggest that maternal higher intake of dairy fat during perinatal period could be potentially associated with offspring’s better cognitive development. These pioneering results call for external validation to challenge their causality.

## Introduction

Maternal suboptimal nutritional intake during pregnancy can influence fetal development and the offspring’s future health (1). Dairy products (DP) are an important source of key nutrients for meeting adequately maternal and fetal nutritional requirements (2). In Western diet, DP contribute importantly to dietary fat intake (3, 4). During perinatal period, maternal fat intake is the main supply of fatty acids for the developing offspring in-utero but also for breastfed infants. Previous studies showed that during breastfeeding period, maternal higher consumption of high-fat content DP was associated with lower linoleic acid (LA) in breast milk (5–8). Interestingly, lower LA level in breast milk was associated with offspring’s greater cognitive development (9). LA is an essential fatty acid but once it is brought in an excessive amount, it could ultimately contribute to an unhealthy n-6:n-3 long-chain polyunsaturated fatty acids ratio (n-6:n-3 LC-PUFAs ratio). However, if and to which extent maternal greater consumption of high-fat content DP, influence offspring’s neurodevelopment, has not yet been investigated to our knowledge.

Pentadecanoic (C15:0) and heptadecanoic acids (C17:0) have been suggested as biomarkers of dairy fat consumption (10). Indeed, based on a recent meta-analysis on observational studies, dairy fat consumption was consistently correlated with C15:0 and to a lesser extent with C17:0 levels in plasma or serum (10). Specifically, these two odd-chain saturated fatty acids (OCFA) originate from rumen fermentation and are then mobilized for the production of milk fat in the mammary gland (11). From our previous findings, C15:0 and C17:0 levels were positively associated with maternal consumption of high-fat content DP, in multiple perinatal biofluids namely maternal and cord red blood cells (RBC) membrane and colostrum (5). These results support the potential placental and breast milk transfers of C15:0 and C17:0 and their relevance as perinatal biomarkers of maternal dairy fat intake. These hypotheses are substantiated by recently reported cross generational associations between maternal plasmatic OCFAs levels and greater birthweight (12). Assessing maternal dairy fat intake using levels of these two OCFAs in multiple perinatal biofluids could overcome reporting bias inherent to self-reported food questionnaire.

Using data from a French mother-child cohort study, we aimed to investigate the influence of maternal dairy fat intake, assessed using C15:0 and C17:0 levels in several perinatal biofluids, on the offspring’s cognitive development in early childhood. We hypothesized that maternal higher dairy fat intake during breastfeeding period and eventually during pregnancy could be beneficial for the offspring’s cognitive development.

## Materials and Methods

### Study population

Data for this study were retrieved from the EDEN (“Étude des Déterminants pre- et post-natals de la santé de l’ENfant”) mother-child cohort (13). Between 2003 and 2006, 2,002 pregnant women from two maternity units in two-middle size French cities (Nancy and Poitiers) were enrolled during a prenatal clinic visit occurring in their first trimester. Exclusion criteria were French illiteracy, known diabetes before pregnancy, multiple pregnancies or planning to move out of the region within the next 3 years. **Supplemental Figure 1** details the flow chart of the present study that involved only mothers who provided perinatal biological samples with a valid fatty acid composition (maternal blood n=1,875, cord blood n=1,422, colostrum n=934), and whom children underwent any cognitive assessment at ages 2, 3 or 5-6 years follow-up (n=1,082-1,397, n=821-1,060, n=547-710, respectively).

### C15:0 and C17:0 levels in perinatal biofluids

Levels of C15:0 and C17:0 (%total fatty acids) were assessed in maternal and cord RBC membrane and colostrum. Maternal and cord blood samples were collected at 24-28 gestational weeks (GW) and at delivery, respectively. RBCs were separated from plasma by centrifugation and stored at −80°C until analysis. Colostrum samples (volume≈5 mL) were collected from one feed by manual expression within the first week after delivery during maternal stay in the maternity ward and were stored at −80°C until analysis. The fatty acid composition of the RBC membranes and of the colostrum samples were assessed by gas chromatography following a standard protocol (14). Briefly, fatty acid methyl-esters (FAME) were first obtained from a direct methylation procedure on 50 µL of RBC membranes or 100 µL of colostrum samples at 100°C for 1 hour. Using a fused silica capillary fast column (BPX 70, Sigma-Supelco, Saint-Louis, Missouri, USA), FAME were thereafter analyzed by gas chromatography (PerkinElmer Clarus (Shelton, Connecticut, USA), flame ionization detector, Totalchrom software, hydrogen as gas carrier).

### Child cognitive assessment

Child’s cognitive outcomes were assessed at multiple ages and included evaluation of language abilities (15–19) and overall cognition (20, 21). Assessment methods and further details on the outcomes are described in **Table 1**.

**Table 1.**
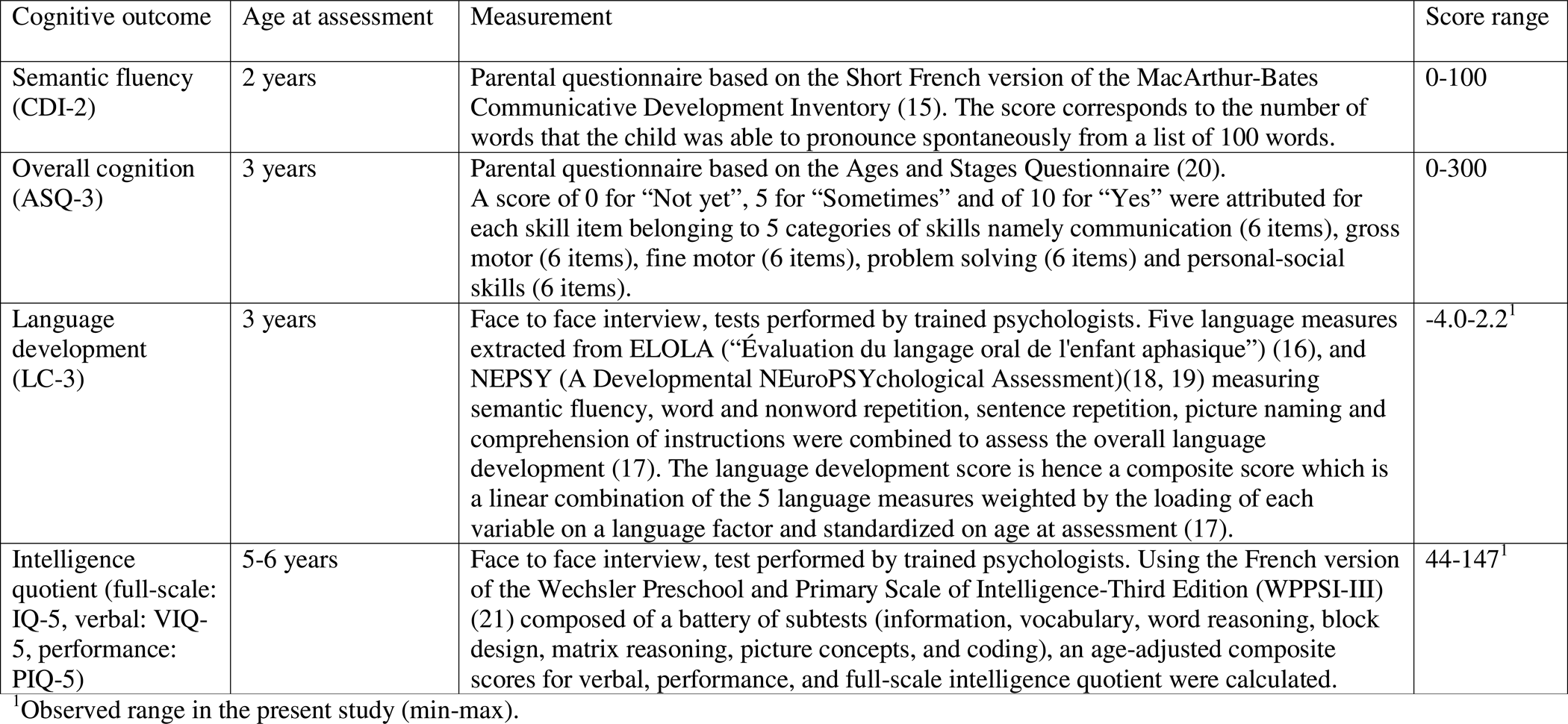
Summary description of the cognitive outcomes.

| Cognitive outcome | Age at assessment | Measurement | Score range |
| --- | --- | --- | --- |
| Semantic fluency (CDI-2) | 2 years | Parental questionnaire based on the Short French version of the MacArthur-Bates Communicative Development Inventory (15). The score corresponds to the number of words that the child was able to pronounce spontaneously from a list of 100 words. | 0-100 |
| Overall cognition (ASQ-3) | 3 years | Parental questionnaire based on the Ages and Stages Questionnaire (20). A score of 0 for “Not yet”, 5 for “Sometimes” and of 10 for “Yes” were attributed for each skill item belonging to 5 categories of skills namely communication (6 items), gross motor (6 items), fine motor (6 items), problem solving (6 items) and personal-social skills (6 items). | 0-300 |
| Language development (LC-3) | 3 years | Face to face interview, tests performed by trained psychologists. Five language measures extracted from ELOLA (“Évaluation du langage oral de l'enfant aphasique”) (16), and NEPSY (A Developmental NEuroPSYchological Assessment)(18, 19) measuring semantic fluency, word and nonword repetition, sentence repetition, picture naming and comprehension of instructions were combined to assess the overall language development (17). The language development score is hence a composite score which is a linear combination of the 5 language measures weighted by the loading of each variable on a language factor and standardized on age at assessment (17). | -4.0-2.2<sup>1</sup> |
| Intelligence quotient (full-scale: IQ-5, verbal: VIQ-5, performance: PIQ-5) | 5-6 years | Face to face interview, test performed by trained psychologists. Using the French version of the Wechsler Preschool and Primary Scale of Intelligence-Third Edition (WPPSI-III) (21) composed of a battery of subtests (information, vocabulary, word reasoning, block design, matrix reasoning, picture concepts, and coding), an age-adjusted composite scores for verbal, performance, and full-scale intelligence quotient were calculated. | 44-147<sup>1</sup> |

<sup>1</sup>Observed range in the present study (min-max).

### Covariates

At 24-28 GW, information about maternal characteristics (age at delivery, education attainment, smoking and alcohol intake during pregnancy), household income and paternal education attainment were collected during a face-to-face interview. Maternal pre-pregnancy body mass index (BMI) was derived using self-reported pre-pregnancy weight and measured height. Maternal depression score during pregnancy was assessed by a self-administered questionnaire (22).

At delivery, maternal diet over the last trimester of pregnancy was assessed using a self-administered validated food frequency questionnaire (23). Using principal component analysis, a “healthy” component or pattern characterized by a greater consumption of fruits and vegetables, high-fat content DP, legumes, lean fish and whole grain cereals was identified (24). Each mother coordinate on the “healthy” component was defined as her “healthy” diet score.

Gestational age at delivery was determined from the date of the last menstrual period and early standard ultrasound fetal measurement. Any breastfeeding was defined as receiving any breast milk (whether exclusively or partially) and its duration was retrieved from questionnaires at 4 months, 8 months, and 1 and 2 years completed by mothers.

At age 2 years, parents reported the child main caregiver during daytime (parents, family/nursery or nanny) and the frequencies of storytelling, singing and playing (never/ less than 1 per week, 1-2 times per week, 3-5 times per week, 7 times per week). An index of the weekly frequency of any caregiver activities with the child was calculated by averaging the aforementioned frequencies of stimulation activities. The same index was built at age 3 years.

At age 3 years, mothers also reported whether the child attended preschool. In France, preschool is a public service and can start from 2.5 years of age depending on the child birth month for a given calendar year. For each child, the time interval between the cognitive assessment at age 3 years and the preschool starting date was calculated (“preschool attendance”).

At age 5-6 years follow-up visit, trained research staff administered a questionnaire of 21 items measuring 3 subscales (25), e.g., language stimulation, learning stimulation and variety in experience. From this questionnaire, a family stimulation score was derived.

### Statistical analysis

Characteristics of the included and non-included participants were described using means and frequencies. C15:0 and C17:0 levels were standardized (z-scores, SD) to facilitate comparison of their association estimates. For each perinatal biofluid, associations of C15:0 and C17:0 levels with child’s cognitive scores were studied using simple and multiple linear regressions models. The linearity of the associations was assessed by testing the associations between C15:0 and C17:0 levels quintiles and cognitive outcomes for one quintile increase. Models (M1) were adjusted for study center, maternal age at delivery, child’s sex and age at cognitive assessment (for cognitive outcomes non-standardized on age), family socioeconomic context and maternal health and lifestyle: maternal pre-pregnancy BMI, parity, tobacco and alcohol consumption during pregnancy, depression score during pregnancy, monthly household income, parental education attainment, child’s main caregiver at age 2 years, stimulation activities level at age at cognitive assessment and preschool attendance (in models with outcomes at age 3 years only). To attempt to disentangle the respective influence of maternal high-fat content DP consumption and maternal balanced diet on children neurodevelopment, we further adjusted the models for maternal healthy diet score (M2) (24, 26). Because we hypothesized that n-6:n-3 LC-PUFAs ratio, could be a potential mediator of the studied associations, we did not perform any adjustment on it. Colostrum models were additionally adjusted for gestational age, the day of colostrum samples collection and any breastfeeding duration.

To better understand whether breastfeeding duration moderates the associations between C15:0 and C17:0 levels in colostrum with cognitive outcomes, the interaction was tested. Even if preterm birth (<37 GW) is associated with lower cognitive outcomes later on, they represented less than 7% of our study sample. Hence, we believe that they would have little influence on our findings and removed them only in a sensitivity analysis. We also further removed children with an IQ lower than 70 (“extremely low”, ≈1-2%), as they may have underlying health conditions. Missing data on covariates were handled with multiple imputation by chained equation under the assumption of missing at random (20 datasets) (27). Significance level was set at alpha=0.05 for all tests except for interaction test (alpha=0.20). A complete case analysis was run to evaluate the influence of the imputation on our findings.

Analyses were performed using SAS (version 9.4; SAS Institute, Cary, NC, USA) and forests plots were obtained using R software (version 4.1.1; R Core Team (2017). R: A language and environment for statistical computing. R Foundation for Statistical Computing, Vienna, Austria. URL https://www.R-project.org/.). The analysis plan was pre-registered online on OSF (28).

## Results

Characteristics of the study population with available data on both the biofluids samples and the IQ score at age 5-6 years were described in comparison with EDEN full cohort in **Table 2**. Briefly, EDEN full cohort is characterized by a maternal mean age of 29 (± 5) years at delivery and mothers with a high educational attainment (54% of mothers with at least a 2-year university degree). About a quarter of mothers were overweight prior to pregnancy, smoked during pregnancy or never breastfed. Compared with EDEN full cohort, participants still followed at age 5 years with at least one biofluid sample collected prenatally and an IQ-5 score, displayed lower levels of maternal depression and tobacco consumption during pregnancy and a better socioeconomic status in terms of educational attainment and income. Specifically, mothers who provided colostrum samples had an even higher socioeconomic status and longer breastfeeding duration.

**Table 2.**
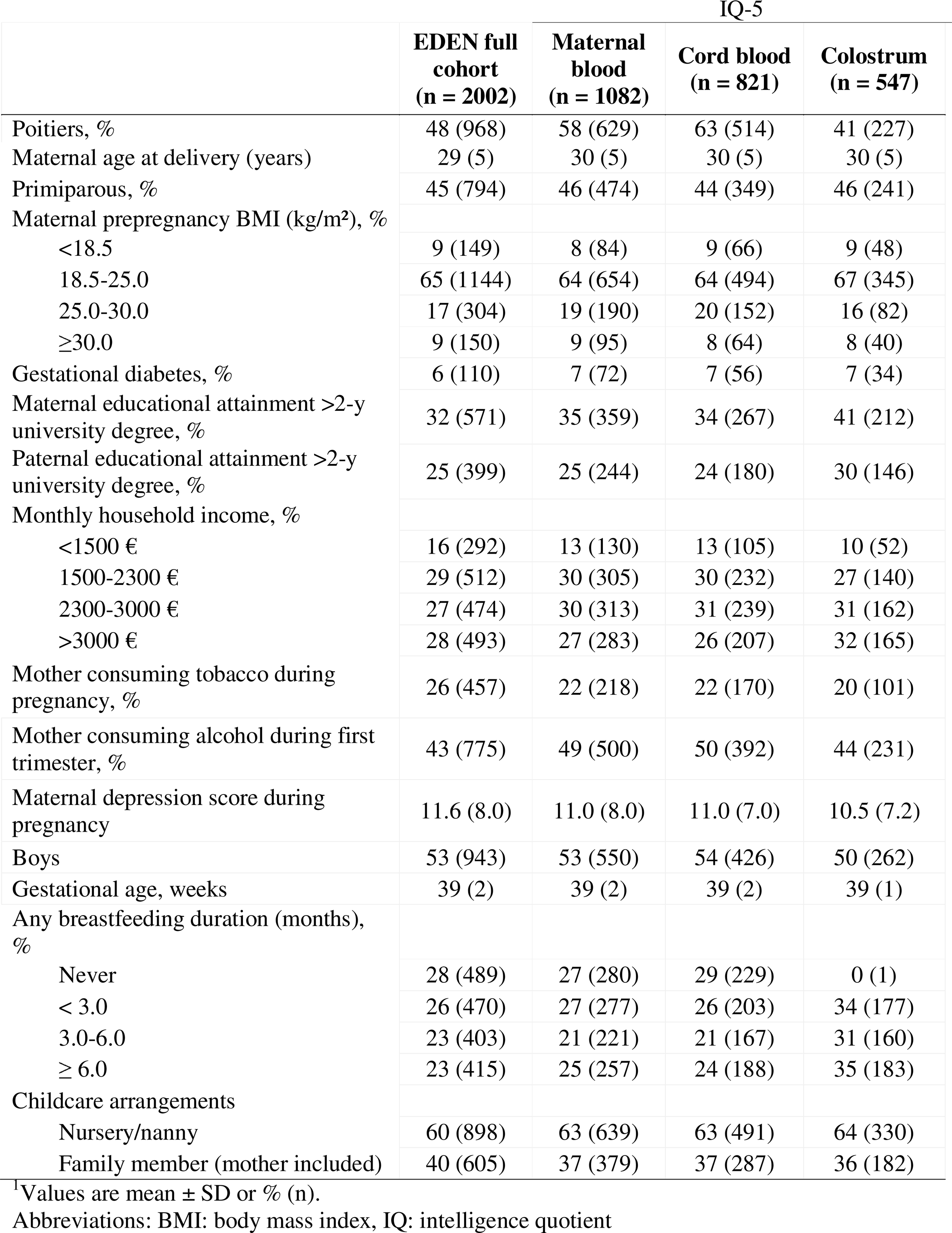
Characteristics of EDEN full cohort participants and of participants with both biofluids samples and IQ-5 score at age 5-6 years^1^.

Overall, differences observed between study populations with cognitive outcomes assessed at ages 2 or 3 years and EDEN full cohort were of similar magnitude of those aforementioned at age 5-6 years (**Supplemental Table 1 and 2)**.

Across study populations, no differences in C15:0 and C17:0 levels were observed and minor ones (up to 2.4% difference) in LA level and n-6:n-3 LC-PUFAs ratio were noticeable (**Supplemental Table 3).**

Associations between C15:0 and C17:0 levels in each biofluid and children language abilities at ages 2 or 3 years (CDI-2 and LC-3, respectively) are presented in **Figure 1**. Only adjusted associations are described. Overall, all associations with either C15:0 or C17:0 remained similar after accounting for maternal healthy diet score.

**Figure 1.**
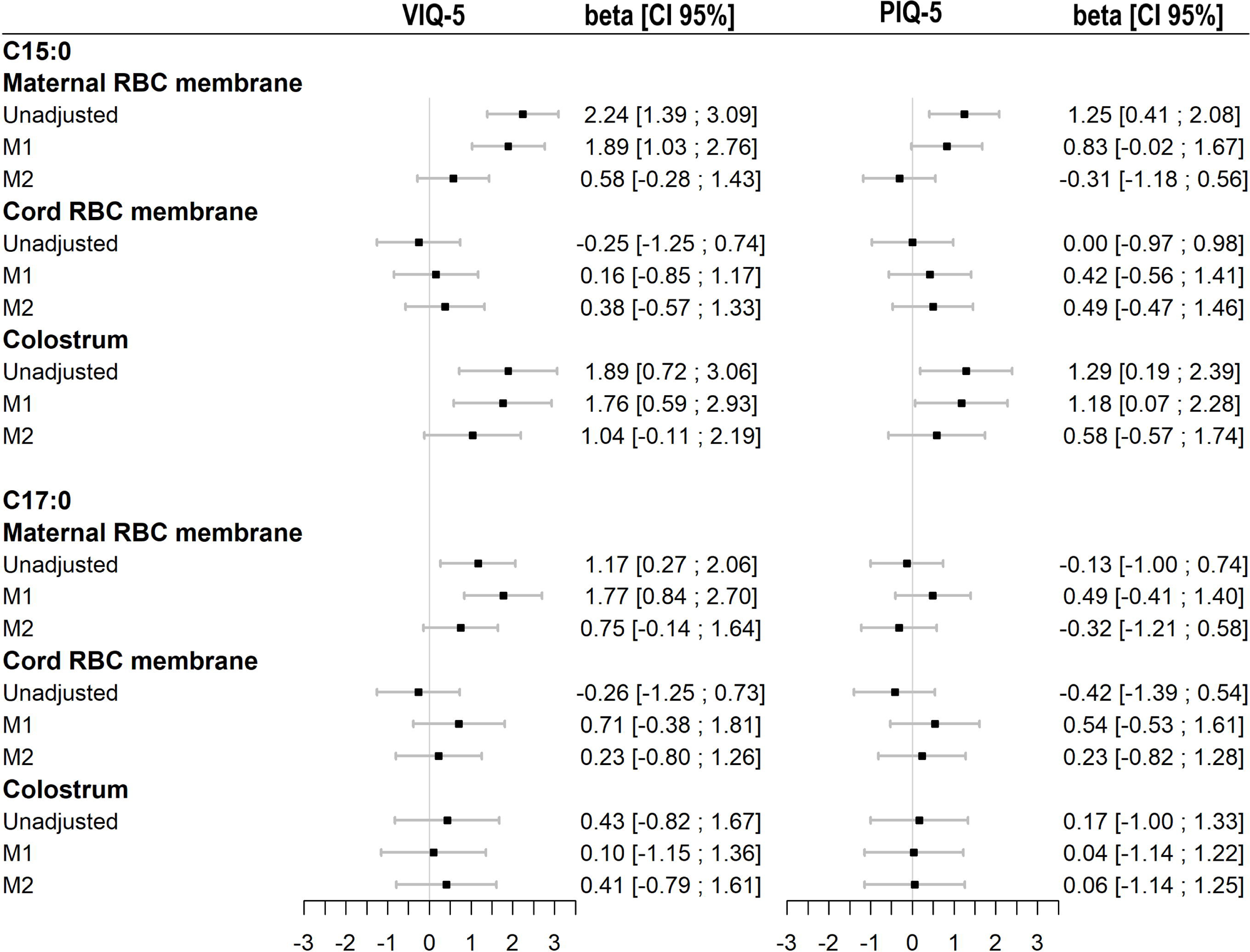
Associations between C15:0 and C17:0 levels in perinatal biofluids and child’s language abilities at ages 2 (CDI-2; on the left) or 3 years (LC-3; on the right). for CDI-2, linear regression models were run on n=1397, n=1060 and n=710 for maternal RBC, cord RBC membranes and colostrum, respectively. For LC-3, linear regression models were run on n=1220, n=927 and n=621 for maternal, cord RBC membranes and colostrum, respectively. Values are β [CI95%] expressed as 1 point (CDI-2) or 1SD (LC-3) change in cognitive outcomes per 1SD increase in C15:0 or C17:0 levels (standardized for comparison). M1 was adjusted for study center, maternal age at delivery, child’s sex, age at cognitive assessment (only in models with CDI-2), maternal pre-pregnancy BMI, parity, tobacco and alcohol consumption during pregnancy, depression score during pregnancy, monthly household income, parental education attainment, child main caregiver at age 2 years, stimulation activities level at age at cognitive assessment, preschool attendance (only in models with LC-3) and M2 was further adjusted for maternal healthy diet score. Models for colostrum, were additionally adjusted for gestational age, the day of collection and any breastfeeding duration.

No association was found with C15:0 level in any biofluids and CDI-2. Higher C15:0 level in maternal RBC membrane and in colostrum was associated with higher LC-3. After stratifying on any breastfeeding duration, C15:0 level in colostrum was positively associated with LC-3 with an increasing magnitude following prolonged breastfeeding duration (P_interaction_=0.09, **Table 3**).

**Table 3.**
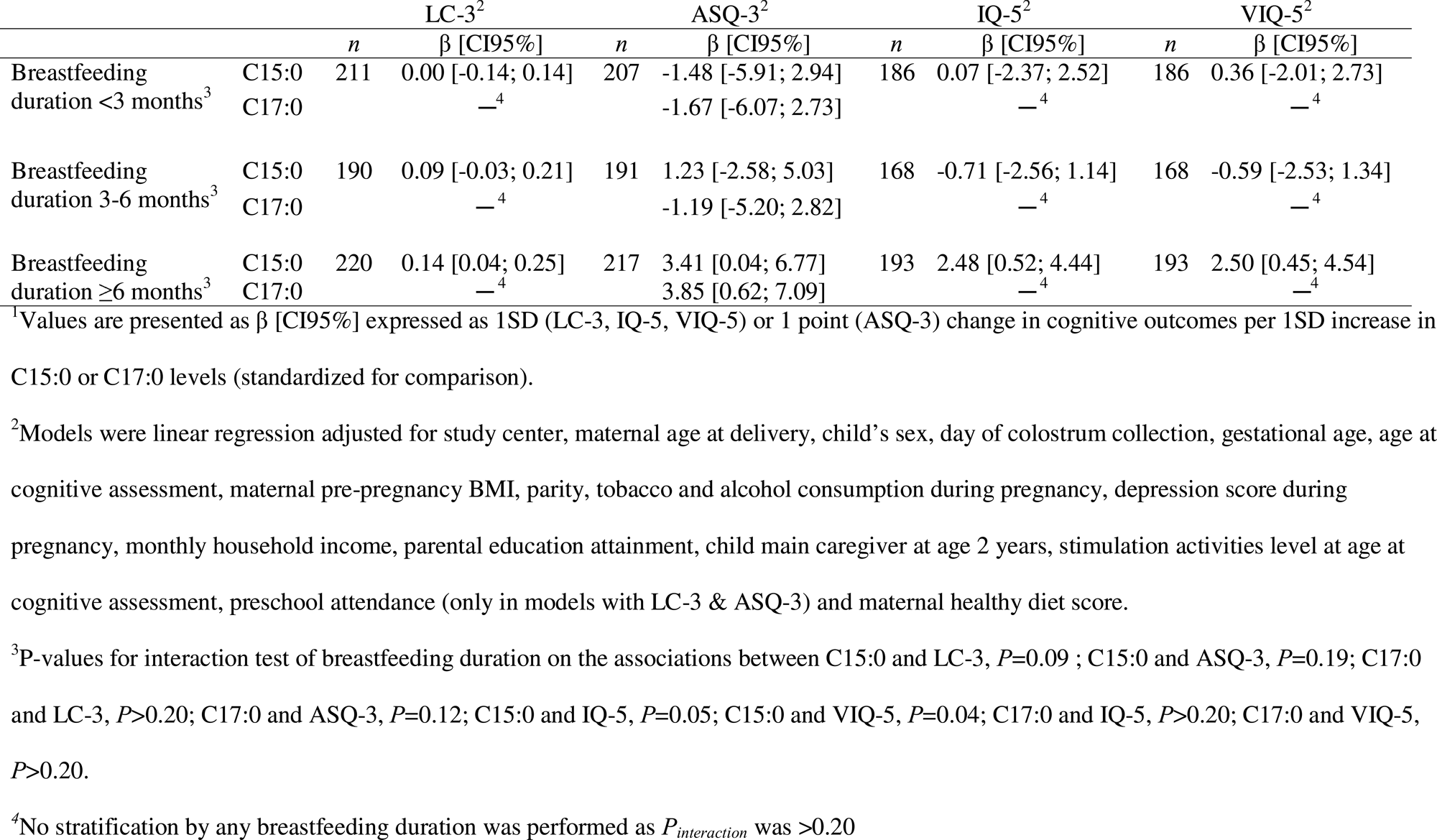
Associations between C15:0 and C17:0 level in colostrum with LC-3, ASQ-3, IQ-5 and VIQ-5 respectively, stratified by any breastfeeding duration^1^.

Higher C17:0 level in cord RBC membrane was associated with higher CDI-2 and LC-3; a similar trend was observed for C17:0 level in maternal RBC. No association was found between C17:0 level in colostrum and either CDI-2 or LC-3.

Associations between DP biomarkers (C15:0 and C17:0) in each biofluid and child’s overall cognition scores at ages 3 years and 5-6 years (ASQ-3 and IQ-5, respectively) are presented in **Figure 2**. Further results on VIQ-5 and PIQ-5 are detailed in **Figure 3**. No associations were observed with either C15:0 or C17:0 and overall cognitive scores at any ages. Once we stratified on breastfeeding duration, marginal positive associations between C15:0 level in colostrum and ASQ-3, IQ-5 and VIQ-5 and between C17:0 level in colostrum and ASQ-3, were observed for children breastfed at least 6 months (P_interaction_=0.19, 0.05, 0.04 and 0.12 respectively, Table 3).

**Figure 2.**
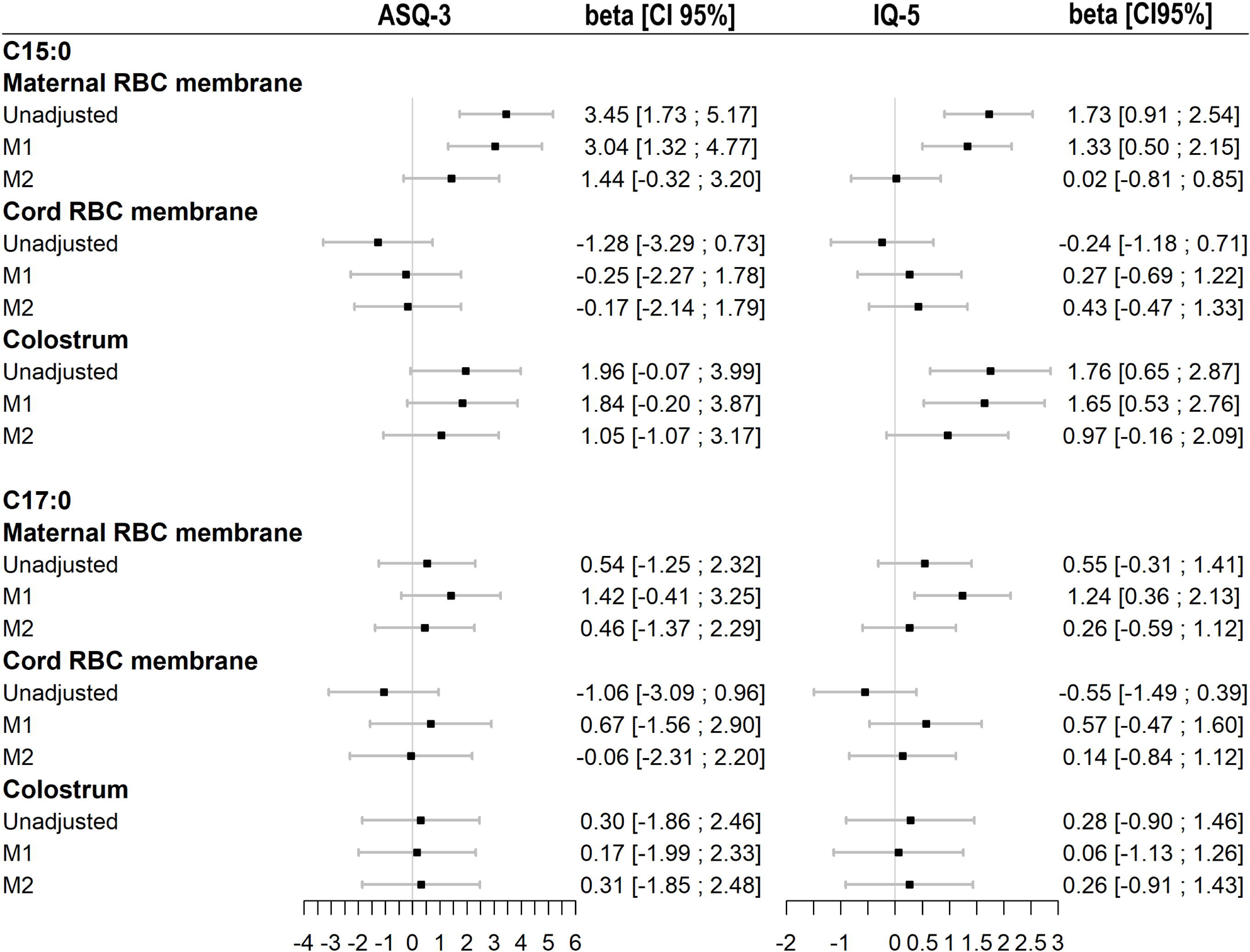
Associations between C15:0 and C17:0 levels in perinatal biofluids and child’s overall cognitive development scores at ages 3 (ASQ-3; on the left) or 5-6 years (IQ-5; on the right). For ASQ-3, linear regression models were run on n=1207, n=929 and n=615 for maternal RBC, cord RBC membranes and colostrum, respectively. For IQ-5, linear regression models were run on n=1082, n=821and n=547 for maternal, cord RBC membranes and colostrum, respectively. Values are β [CI95%] expressed as 1 point (ASQ-3) or 1SD (IQ-5) change in cognitive outcomes per 1SD increase in C15:0 or C17:0 levels (standardized for comparison). M1 was adjusted for study center, maternal age at delivery, child’s sex, age at cognitive assessment (only in models with ASQ-3), maternal pre-pregnancy BMI, parity, tobacco and alcohol consumption during pregnancy, depression score during pregnancy, monthly household income, parental education attainment, child main caregiver at age 2 years, stimulation activities level at age at cognitive assessment, preschool attendance (only in models with ASQ-3) and M2 was further adjusted for maternal healthy diet score. Models for colostrum, were additionally adjusted for gestational age, the day of collection and any breastfeeding duration

**Figure 3.**
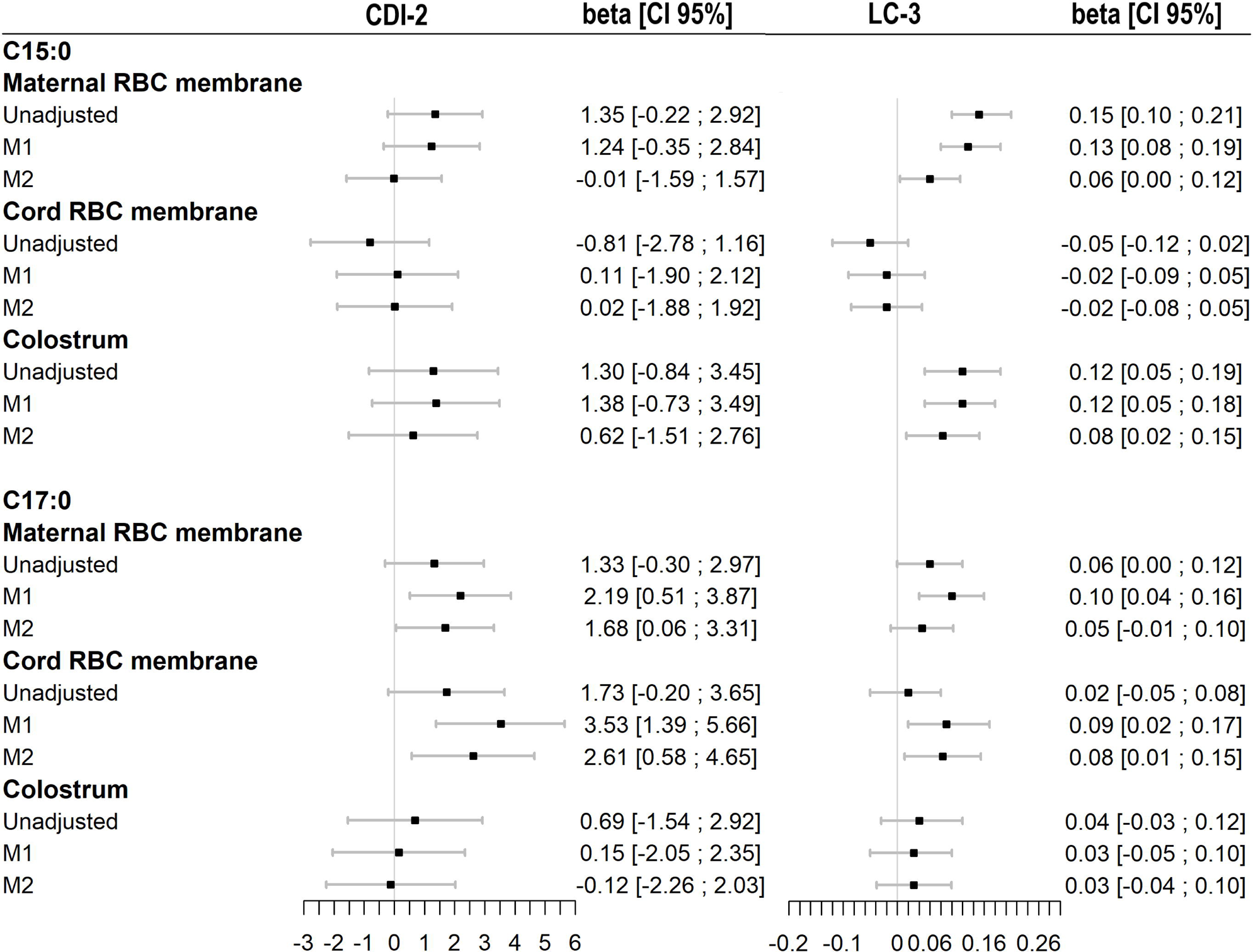
Associations between C15:0 and C17:0 levels in perinatal biofluids with VIQ-5 (on the left) and PIQ-5 at age 5-6 years (on the right). For both cognitive outcomes, linear regressions models were run on n=1082, n=821 and n=541 for maternal, cord RBC membranes and colostrum, respectively. Values are β [CI95%] expressed as 1SD change in PIQ-5/VIQ-5 per 1SD increase in C15:0 or C17:0 levels (standardized for comparison). M1 was adjusted for study center, maternal age at delivery, child’s sex, maternal pre-pregnancy BMI, parity, tobacco and alcohol consumption during pregnancy, depression score during pregnancy, monthly household income, parental education attainment, child main caregiver at age 2 years, stimulation activities level at age 5-6 years and M2 was further adjusted for maternal healthy diet score. Models for colostrum, were additionally adjusted for gestational age, the day of collection and any breastfeeding duration

Across all studied associations, results remained similar after removing preterm children, children with extremely low IQ and children with missing data for covariates from our analysis (**Supplemental Figure 2, Figure 3, Figure 4, Table 4, Table 5, Table 6**).

## Discussion

From our results, C15:0 and C17:0 levels in maternal RBC membrane were positively associated with early language scores. In colostrum, C15:0 level was more consistently associated with early language scores than C17:0 level. In contrast, in cord RBC membrane, most associations were observed with C17:0 level. In children breastfed at least 6 months, the early positive associations with dairy fat biomarkers in colostrum were also observed at ages 3 and 5-6 years with overall cognitive development scores and verbal IQ

Currently, we did not identify any other study investigating the link between either pregnant or breastfeeding women’s dairy fat consumption and their offspring’s cognitive outcomes. Yet, it is now widely recognized that maternal diet plays a critical role on fetal growth including brain development (1). Recent evidence suggests that maternal greater plasmatic levels of OCFAs (C15:0 and C17:0 in particular) during pregnancy were associated with greater birthweight (12). From our previous findings, C15:0 and C17:0 levels in perinatal biofluids (maternal and cord RBC membranes, colostrum) were positively associated with maternal high-fat content DP consumption during pregnancy, especially cheese consumption This might suggest a potential transfer of these biomarkers to the fetus and breastfed infants through placenta and breast milk, respectively.

Here, our results further suggest positive associations between these dairy fat biomarkers and the child’s early language development. Importantly, these associations persisted even after accounting for maternal healthy dietary pattern scores during pregnancy. This indicates that the potential benefits of high-fat content DP consumption on child’s cognitive outcomes may not be solely explained by maternal healthier diet (24). Interestingly, child’s overall cognition (ASQ-3, IQ-5) was only associated with dairy fat biomarkers contained in colostrum, particularly in children breastfed at least 6 months. One potential explanation of the association between higher levels of dairy fat biomarkers in colostrum and children’s better cognitive outcomes is the lower LA level in breast milk, which itself could contribute to a more balanced n-6:n-3 LC-PUFAs ratio. Indeed, within the same cohort study, lower LA level in colostrum was associated with both higher maternal high-fat content DP consumption and children’s better cognitive outcomes (motor skills at age 2 years and ASQ-3) (5, 9). Furthermore, hypotheses were made on the direct involvement of C15:0 and C17:0 in the biosynthesis of odd-numbered very long chain fatty acids for the glycosphingolipids of the nervous tissue (29). Other nutritional components of DP may also explain our findings. DP are an important source of key nutrients that were related to cognitive function such as essential amino acids, vitamins (B1, B2, B12, D) and phosphorus, potassium, iodine, calcium (30) and polar lipids (31). Other DP nutrients were also related to cognitive function mostly in older adults such as bioactive peptides and probiotics (32).

Here, we did not find consistent associations between C15:0 and C17:0 levels across biofluids. We hypothesized that differential association of C15:0 and C17:0 across biofluids with children’s cognitive outcomes may originate from differences in how they are metabolized in the placenta and mammary glands. This is supported by our previous findings showing poor intercorrelation of C15:0 and C17:0 levels in the different perinatal biofluids (except for C15:0 levels, between maternal RBC membrane and colostrum) (5). C15:0 and C17:0 levels in cord RBC membrane and colostrum may be a better reflection of the fetus and breastfed infant actual status respectively, than levels in maternal RBC membrane. However, endogenous synthesis pathways were suggested for C17:0 and less likely for C15:0 (29, 33).

Hence, we acknowledge that further fundamental research is needed to substantiate our hypothesis.

Our study has some limitations. Caution is needed once we compared our findings across ages, as we used different cognitive scores evaluating different aspect of cognitive development and with different assessment methods. However, having different cognitive assessments allows us to have a more comprehensive investigation of the eventual influence of DP biomarkers on children cognitive development. Furthermore, one may argue that across follow-up waves, our findings might be influenced by the attrition bias, inherent to most cohort studies. For instance, children with lower language abilities tend to belong to socioeconomically more disadvantaged families and hence have higher likelihood to being loss to follow up. However, from our descriptive results, the attrition bias towards more socioeconomically advantaged families was observed since age 2 years and since that age, family’s socioeconomic characteristics remained mostly similar across follow-up waves. Overall, our study sample is highly educated which limits the transportability of our findings to lower socioeconomic populations. DP consumption, in particular cheese, is high in the French population which could increase our likelihood to find an association but also limit the replicability of our results. Additionally, as most children attend preschool around age 3 years in France, it might smooth out important language disparities observed in infancy. One of the strengths of our study is the use of dairy fat biomarkers rather than self-reported DP consumption to assess objectively maternal dairy fat consumption. Having these biomarkers in multiple perinatal biofluids collected at different development stages of the offspring allow us to better investigate different biological pathways through which maternal dairy fat consumption could affect children cognitive development.

Our study is the first to explore the association between maternal dairy fat intake during perinatal period and children’s cognitive development. Using C15:0 and C17:0 as biomarkers of maternal dairy fat intake in several perinatal biofluids, we found positive associations with children’s language abilities at preschool ages. Children’s greater language abilities were hence associated with higher C15:0 and C17:0 levels in maternal RBC membrane, C17:0 level in cord RBC membrane and higher C15:0 level in colostrum. These findings trigger interest for investigating maternal high-fat content DP consumption during the first 1,000 days on her offspring’s cognitive outcomes. They need to be corroborated by further studies in order to challenge their causality. To better understand the underlying mechanisms that could explain the observed associations, further fundamental studies are warranted as well to unveil the physiological pathways.

## Data availability Statement

The data underlying the findings cannot be made freely available for ethical and legal restrictions imposed, because this study includes a substantial number of variables that, together, could be used to re-identify the participants based on a few key characteristics and then be used to have access to other personal data. Therefore, the French ethics authority strictly forbids making these data freely available. However, they can be obtained upon request from the EDEN principal investigator. Readers may contact to request the data. The analytic code will be made available upon request pending application and approval.

## Acknowledgments

We are extremely grateful to all the families who took part in this study, the midwives for their help in recruiting them, and the whole EDEN team, which includes psychologists, research scientists, engineers, technicians and managers. The authors thank the EDEN mother-child cohort study group, whose members are I. Annesi-Maesano, J.Y. Bernard, J. Botton, M.A. Charles, P. Dargent-Molina, B. de Lauzon-Guillain, P. Ducimetière, M. de Agostini, B. Foliguet, A. Forhan, X. Fritel, A. Germa, V. Goua, R. Hankard, B. Heude, M. Kaminski, B. Larroque, N. Lelong, J. Lepeule, G. Magnin, L. Marchand, C. Nabet, F. Pierre, R. Slama, M.J. Saurel-Cubizolles, M. Schweitzer, O. Thiebaugeorges.

## Statement of authors’ contributions to manuscript

The authors’ responsibilities were as follows—WLY, JYB and BH designed research. WLY conducted research. MA and CS provided essential materials. WLY performed statistical analysis. WLY wrote the initial draft. JYB, MA, CS, MAC, BH and HP critically reviewed and edited the draft. WLY had primary responsibility for final content. All authors read and approved the final manuscript.

## Ethical approval

The EDEN mother-child cohort study was approved by the ethics committee of Kremlin Bicêtre (CPP reference 02-70, December 2002; CCTIRS reference 02-189, July 2002) and the National Data Protection Authority (“Commission Nationale de l’Informatique et des Libertés”, CNIL reference 902267, December 2002).

