## Supplemental Figure 1 for "Associations between perinatal biomarkers of maternal dairy fat intake and child cognitive development: results from the EDEN mother-child cohort"

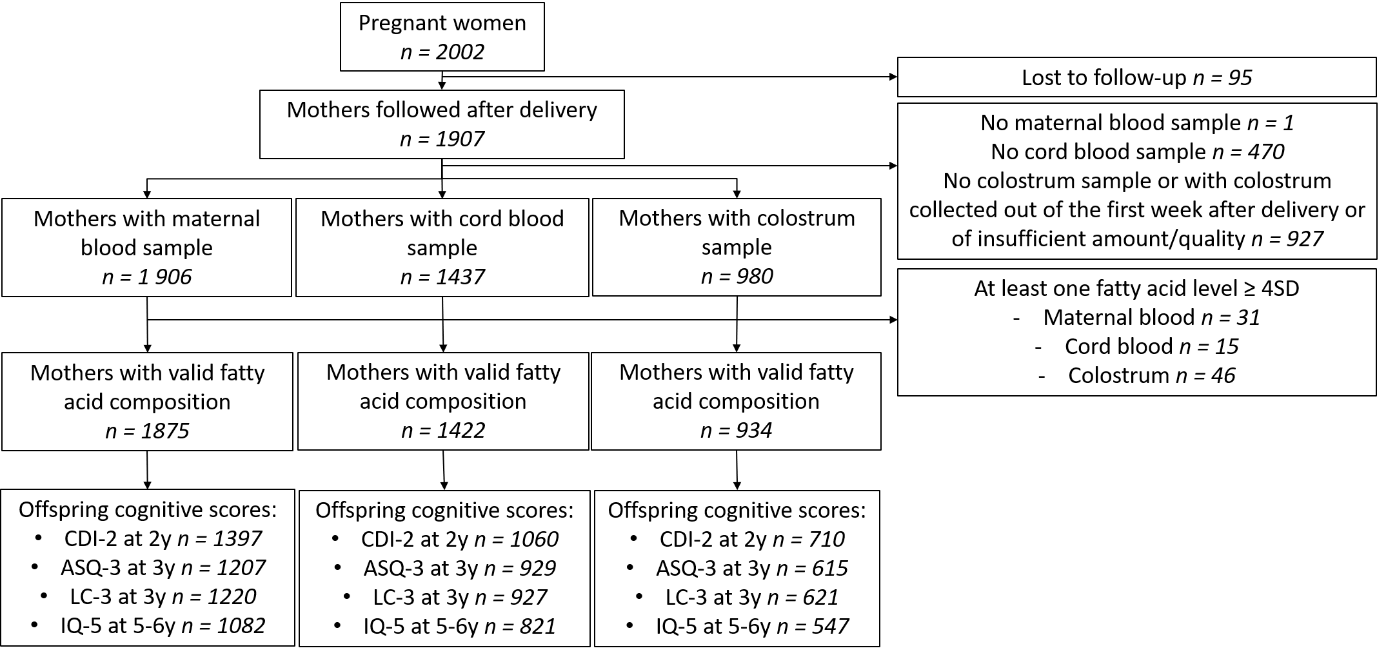

**Supplemental Figure 1**. Study flow chart.

Abbreviations: ASQ-3: Ages and Stages Questionnaire at age 3 years, CDI-2: Communicative Development Inventory at age 2 years, IQ-5: intelligence quotient at age 5-6 years, LC-3: Language composite score at age 3 years
