## Supplemental Table 6 for "Associations between perinatal biomarkers of maternal dairy fat intake and child cognitive development: results from the EDEN mother-child cohort"

Supplemental Table 6. Associations between C15:0 and C17:0 level in colostrum with LC-3, ASQ-3, IQ-5 and VIQ-5, respectively, stratified by any breastfeeding duration with children with non-missing data for covariates^1^

|  |  | LC-3^2^ | | ASQ-3^2^ | | IQ-5^2^ | | VIQ-5^2^ | |
| --- | --- | --- | --- | --- | --- | --- | --- | --- | --- |
|  |  | *n* | β [CI95%] | *n* | β [CI95%] | *n* | β [CI95%] | *n* | β [CI95%] |
| Breastfeeding duration <3 months^3^ | C15:0 | 167 | -0.05 [-0.22; 0.12] | 170 | -1.60 [-6.59; 3.39] | 142 | 0.18 [-2.77; 3.12] | 142 | 0.68 [-2.17; 3.53] |
|  | C17:0 |  | *─*^4^ |  | -1.45 [-6.38; 3.48] |  | *─*^4^ |  | *─*^4^ |
| Breastfeeding duration 3-6 months^3^ | C15:0 | 155 | 0.10 [-0.04; 0.24] | 162 | 0.28 [-4.08; 4.64] | 136 | -0.88 [-3.03; 1.26] | 136 | -1.15 [-3.44; 1.14] |
|  | C17:0 |  | *─*^4^ |  | -0.32 [-4.65; 4.01] |  | *─*^4^ |  | *─*^4^ |
| Breastfeeding duration ≥6 months^3^ | C15:0 | 158 | 0.13 [0.00; 0.26] | 159 | 2.23 [-1.76; 6.22] | 140 | 1.45 [-1.00; 3.89] | 140 | 1.30 [-1.28; 3.88] |
|  | C17:0 |  | *─*^4^ |  | 3.56 [-0.28; 7.41] |  | *─*^4^ |  | *─*^4^ |

^1^Values are presented as β [CI95%] expressed as 1SD (LC-3, IQ-5, VIQ-5) or 1 point (ASQ-3) change in cognitive outcomes per 1SD increase in C15:0 or C17:0 levels (standardized for comparison).

^2^Models were linear regression adjusted for study center, maternal age at delivery, child’s sex, day of colostrum collection, gestational age, age at cognitive assessment (only in models with ASQ-3), maternal pre-pregnancy BMI, parity, tobacco and alcohol consumption during pregnancy, depression score during pregnancy, monthly household income, parental education attainment, child main caregiver at age 2 years, stimulation activities level at age matched with the studied outcome, preschool attendance (only in models with LC-3 & ASQ-3) and maternal healthy diet score.

^3^ P-values for interaction test of breastfeeding duration on the associations between C15:0 and LC-3, *P*=0.09; C15:0 and ASQ-3, *P*=0.19; C17:0 and LC-3, *P*>0.20; C17:0 and ASQ-3, *P*=0.12; C15:0 and IQ-5, *P*=0.05; C15:0 and VIQ-5, *P*=0.04; C17:0 and IQ-5, *P*>0.20; C17:0 and VIQ-5, *P*>0.20.

^4^No stratification by any breastfeeding duration was performed as *P_interaction_* was >0.20.
