## Supplemental Table 5 for "Associations between perinatal biomarkers of maternal dairy fat intake and child cognitive development: results from the EDEN mother-child cohort"

Supplemental Table 5. Associations between C15:0 and C17:0 level in colostrum with LC-3, ASQ-3, IQ-5 and VIQ-5, respectively, stratified by any breastfeeding duration after removing children with an IQ<70^1^

|  |  | LC-3^2^ | | ASQ-3^2^ | | IQ-5^2^ | | VIQ-5^2^ | |
| --- | --- | --- | --- | --- | --- | --- | --- | --- | --- |
|  |  | *n* | β [CI95%] | *n* | β [CI95%] | *n* | β [CI95%] | *n* | β [CI95%] |
| Breastfeeding duration <3 months^3^ | C15:0 | 207 | 0.00 [-0.13; 0.14] | 202 | -1.31 [-5.48; 2.86] | 183 | 0.26 [-1.94; 2.47] | 183 | 0.54 [-1.64; 2.72] |
|  | C17:0 |  | *─*^4^ |  | -1.96 [-6.12; 2.19] |  | *─*^4^ |  | *─*^4^ |
| Breastfeeding duration 3-6 months^3^ | C15:0 | 190 | 0.10 [-0.03; 0.22] | 191 | 1.20 [-2.66; 5.06] | 167 | -1.11 [-2.93; 0.70] | 167 | -0.81 [-2.75; 1.14] |
|  | C17:0 |  | *─*^4^ |  | -0.98 [-3.00; 1.04] |  | *─*^4^ |  | *─*^4^ |
| Breastfeeding duration ≥6 months^3^ | C15:0 | 220 | 0.14 [0.03; 0.25] | 216 | 2.50 [-0.50; 5.51] | 190 | 2.06 [0.36; 3.76] | 190 | 2.11 [0.28; 3.93] |
|  | C17:0 |  | *─*^4^ |  | 1.66 [-0.32; 3.63] |  | *─*^4^ |  | *─*^4^ |

^1^Values are presented as β [CI95%] expressed as 1SD (LC-3, IQ-5, VIQ-5) or 1 point (ASQ-3) change in cognitive outcomes per 1SD increase in C15:0 or C17:0 levels (standardized for comparison).

^2^Models were linear regression adjusted for study center, maternal age at delivery, child’s sex, day of colostrum collection, gestational age, age at cognitive assessment (only in models with ASQ-3), maternal pre-pregnancy BMI, parity, tobacco and alcohol consumption during pregnancy, depression score during pregnancy, monthly household income, parental education attainment, child main caregiver at age 2 years, stimulation activities level at age matched with the studied outcome, preschool attendance (only in models with LC-3 & ASQ-3) and maternal healthy diet score.

^3^ P-values for interaction test of breastfeeding duration on the associations between C15:0 and LC-3, *P*=0.09; C15:0 and ASQ-3, *P*=0.19; C17:0 and LC-3, *P*>0.20; C17:0 and ASQ-3, *P*=0.12; C15:0 and IQ-5, *P*=0.05; C15:0 and VIQ-5, *P*=0.04; C17:0 and IQ-5, *P*>0.20; C17:0 and VIQ-5, *P*>0.20.

^4^No stratification by any breastfeeding duration was performed as *P_interaction_* was >0.20.
