## Supplemental Table 4 for "Associations between perinatal biomarkers of maternal dairy fat intake and child cognitive development: results from the EDEN mother-child cohort"

Supplemental Table 4. Associations between C15:0 and C17:0 level in colostrum with LC-3, ASQ-3, IQ-5 and VIQ-5, respectively, stratified by any breastfeeding duration after removing preterm^1^

|  |  | LC-3^2^ | | ASQ-3^2^ | | IQ-5^2^ | | VIQ-5^2^ | |
| --- | --- | --- | --- | --- | --- | --- | --- | --- | --- |
|  |  | *n* | β [CI95%] | *n* | β [CI95%] | *n* | β [CI95%] | *n* | β [CI95%] |
| Breastfeeding duration <3 months^3^ | C15:0 | 196 | 0.04 [-0.10; 0.19] | 193 | 0.77 [-3.77; 5.31] | 175 | 0.82 [-1.68; 3.31] | 175 | 0.88 [-1.55; 3.32] |
|  | C17:0 |  | *─*^4^ |  | 0.59 [-3.94; 5.12] |  | *─*^4^ |  | *─*^4^ |
| Breastfeeding duration 3-6 month^3^ | C15:0 | 187 | 0.08 [-0.05; 0.20] | 186 | 1.42 [-2.51; 5.34] | 163 | -0.87 [-2.77; 1.04] | 163 | -0.80 [-2.81; 1.21] |
|  | C17:0 |  | *─*^4^ |  | 2.09 [-5.11; 3.07] |  | *─*^4^ |  | *─*^4^ |
| Breastfeeding duration ≥6 months^3^ | C15:0 | 211 | 0.15 [0.04; 0.26] | 210 | 3.39 [-0.02; 6.8] | 185 | 2.36 [0.38; 4.34] | 185 | 2.52 [0.43; 4.61] |
|  | C17:0 |  | *─*^4^ |  | 4.06 [0.81; 7.32] |  | *─*^4^ |  | *─*^4^ |

^1^Values are presented as β [CI95%] expressed as 1SD (LC-3, IQ-5, VIQ-5) or 1 point (ASQ-3) change in cognitive outcomes per 1SD increase in C15:0 or C17:0 levels (standardized for comparison).

^2^Models were linear regression adjusted for study center, maternal age at delivery, child’s sex, day of colostrum collection, gestational age, age at cognitive assessment (only in models with ASQ-3), maternal pre-pregnancy BMI, parity, tobacco and alcohol consumption during pregnancy, depression score during pregnancy, monthly household income, parental education attainment, child main caregiver at age 2 years, stimulation activities level at age at cognitive assessment, preschool attendance (only in models with LC-3 & ASQ-3) and maternal healthy diet score.

^3^P-values for interaction test of breastfeeding duration on the associations between C15:0 and LC-3, *P*=0.09; C15:0 and ASQ-3, *P*=0.19; C17:0 and LC-3, *P*>0.20; C17:0 and ASQ-3, *P*=0.12; C15:0 and IQ-5, *P*=0.05; C15:0 and VIQ-5, *P*=0.04; C17:0 and IQ-5, *P*>0.20; C17:0 and VIQ-5, *P*>0.20.

^4^No stratification by any breastfeeding duration was performed as *P_interaction_* was >0.20.
