## Supplemental Table 3 for "Associations between perinatal biomarkers of maternal dairy fat intake and child cognitive development: results from the EDEN mother-child cohort"

Supplemental Table 3. Description of fatty acids levels (C15:0, C17:0, LA, n-6:n-3 LC-PUFA ratio) by biofluids and by study sample^1^

|  | With bio-samples | With CDI-2 | With LC-3 | With ASQ-3 | With IQ-5 |
| --- | --- | --- | --- | --- | --- |
| *Maternal RBC* | *n=1906* | *n=1397* | *n=1207* | *n=1220* | *n=1082* |
| C15:0, % of total fatty acids | 0.17 ± 0.03 | 0.17 ± 0.03 | 0.17 ± 0.03 | 0.17 ± 0.03 | 0.17 ± 0.03 |
| C17:0, % of total fatty acids | 0.31 ± 0.04 | 0.31 ± 0.04 | 0.31 ± 0.04 | 0.31 ± 0.04 | 0.31 ± 0.04 |
| LA, % of total fatty acids | 7.16 ± 0.82 | 7.12 ± 0.80 | 7.11 ± 0.80 | 7.10 ± 0.80 | 7.10 ± 0.81 |
| n-6:n-3 LC-PUFA ratio | 2.47 ± 0.50 | 2.43 ± 0.48 | 2.41 ± 0.46 | 2.41 ± 0.46 | 2.41 ± 0.46 |
| *Cord RBC* | *n=1437* | *n=1060* | *n=929* | *n=927* | *n=821* |
| C15:0, % of total fatty acids | 0.12 ± 0.04 | 0.12 ± 0.04 | 0.12 ± 0.04 | 0.12 ± 0.04 | 0.12 ± 0.04 |
| C17:0, % of total fatty acids | 0.32 ± 0.08 | 0.32 ± 0.07 | 0.33 ± 0.08 | 0.33 ± 0.07 | 0.33 ± 0.08 |
| LA, % of total fatty acids | 2.91 ± 0.54 | 2.88 ± 0.42 | 2.89 ± 0.42 | 2.90 ± 0.42 | 2.89 ± 0.43 |
| n-6:n-3 LC-PUFA ratio | 3.41 ± 0.62 | 3.38 ± 0.59 | 3.34 ± 0.58 | 3.35 ± 0.57 | 3.33 ± 0.57 |
| *Colostrum* | *n=980* | *n=710* | *n=615* | *n=621* | *n=547* |
| C15:0, % of total fatty acids | 0.42 ± 0.07 | 0.42 ± 0.07 | 0.42 ± 0.07 | 0.42 ± 0.07 | 0.42 ± 0.07 |
| C17:0, % of total fatty acids | 0.28 ± 0.05 | 0.28 ± 0.05 | 0.28 ± 0.05 | 0.28 ± 0.05 | 0.28 ± 0.05 |
| LA, % of total fatty acids | 9.87 ± 1.90 | 9.75 ± 1.77 | 9.81 ± 1.79 | 9.85 ± 1.80 | 9.80 ± 1.85 |
| n-6:n-3 LC-PUFAs ratio | 1.80 ± 0.42 | 1.79 ± 0.39 | 1.77 ± 0.38 | 1.77 ± 0.37 | 1.77 ± 0.38 |

***^1^***Values are mean ± SD.

Abbreviations: ASQ-3: Ages and Stages Questionnaire at age 3 years, CDI-2: communicative development inventory at age 2 years, IQ-5: Intelligence quotient at age 5-6 years, LA: linoleic acid, LC-3: language composite score at age 3 years, n-6:n-3 LC-PUFA ratio: n-6:n-3 long-chain polyunsaturated fatty acids ratio
