## Supplemental Table 2 for "Associations between perinatal biomarkers of maternal dairy fat intake and child cognitive development: results from the EDEN mother-child cohort"

**Supplemental Table 2**. Characteristics of EDEN full cohort participants and of participants with biofluids samples and LC-3 score at age 3 years^1^

|  |  | LC-3 | | |
| --- | --- | --- | --- | --- |
|  | **EDEN full cohort**  **(n = 2002)** | **Maternal blood (n = 1156)** | **Cord blood**  **(n = 892)** | **Colostrum (n = 588)** |
| Poitiers, % | 48 (968) | 55 (665) | 58 (535) | 38 (238) |
| Maternal age at delivery (years) | 29 (5) | 30 (5) | 30 (5) | 30 (5) |
| Primiparous, % | 45 (794) | 47 (546) | 46 (411) | 48 (283) |
| Maternal pre pregnancy BMI (kg/m²), % |  |  |  |  |
| <18.5 | 9 (149) | 7 (83) | 8 (66) | 8 (47) |
| 18.5-25.0 | 65 (1144) | 66 (756) | 67 (586) | 71 (409) |
| 25.0-30.0 | 17 (304) | 18 (205) | 18 (158) | 15 (85) |
| ≥30.0 | 9 (150) | 8 (94) | 8 (66) | 7 (39) |
| Gestational diabetes, % | 6 (110) | 7 (79) | 7 (61) | 6 (38) |
| Maternal educational attainment >2-y university degree, % | 32 (571) | 38 (434) | 38 (333) | 43 (254) |
| Paternal educational attainment >2-y university degree, % |  |  |  |  |
| Monthly household income, % |  |  |  |  |
| <1500 € | 16 (292) | 12 (133) | 11 (101) | 9 (53) |
| 1500-2300 € | 29 (512) | 29 (335) | 30 (263) | 27 (156) |
| 2300-3000 € | 27 (474) | 29 (336) | 30 (263) | 30 (176) |
| >3000 € | 28 (493) | 30 (347) | 29 (260) | 34 (200) |
| Mother consuming tobacco during pregnancy, % | 26 (457) | 22 (245) | 22 (188) | 20 (117) |
| Mother consuming alcohol during first trimester, % | 43 (775) | 45 (519) | 46 (410) | 42 (248) |
| Maternal depression score during pregnancy | 11.6 (8.0) | 10.8 (7.4) | 10.7 (7.1) | 10.4 (6.9) |
| Boys | 53 (943) | 53 (609) | 54 (483) | 51 (301) |
| Gestational age, weeks | 39 (2) | 39 (2) | 39 (2) | 39 (1) |
| Any breastfeeding duration (months), % |  |  |  |  |
| Never | 28 (489) | 27 (311) | 28 (252) | 0 (1) |
| < 3.0 | 26 (470) | 26 (306) | 26 (236) | 33 (196) |
| 3.0-6.0 | 23 (403) | 22 (251) | 21 (189) | 32 (185) |
| ≥ 6.0 | 23 (415) | 25 (287) | 24 (214) | 35 (205) |
| Childcare arrangements |  |  |  |  |
| Nursery/nanny | 60 (898) | 62 (719) | 62 (557) | 64 (376) |
| Family member (mother included) | 40 (605) | 38 (437) | 38 (335) | 36 (212) |

^1^ Values are mean ± SD or % (n).

Abbreviations: BMI: body mass index, LC: language composite score
