## Supplemental Table 1 for "Associations between perinatal biomarkers of maternal dairy fat intake and child cognitive development: results from the EDEN mother-child cohort"

**Supplemental Table 1**. Characteristics of EDEN full cohort participants and of participants with biofluids samples and CDI-2 score at age 2 years^1^

|  |  | CDI-2 | | |
| --- | --- | --- | --- | --- |
|  | **EDEN full cohort**  **(n = 2002)** | **Maternal blood (n = 1341)** | **Cord blood**  **(n = 1021)** | **Colostrum (n = 677)** |
| Poitiers, % | 48 (968) | 48 (672) | 52 (547) | 32 (226) |
| Maternal age at delivery (years) | 29 (5) | 30 (5) | 30 (5) | 30 (5) |
| Primiparous, % | 45 (794) | 47 (634) | 46 (473) | 48 (324) |
| Maternal pre pregnancy BMI (kg/m²), % |  |  |  |  |
| <18.5 | 9 (149) | 8 (105) | 8 (83) | 9 (58) |
| 18.5-25.0 | 65 (1144) | 67 (886) | 68 (677) | 72 (481) |
| 25.0-30.0 | 17 (304) | 17 (225) | 17 (171) | 14 (91) |
| ≥30.0 | 9 (150) | 8 (104) | 7 (70) | 6 (40) |
| Gestational diabetes, % | 6 (110) | 6 (84) | 6 (64) | 5 (37) |
| Maternal educational attainment >2-y university degree, % | 32 (571) | 37 (488) | 36 (370) | 43 (289) |
| Paternal educational attainment >2-y university degree, % | 25 (399) | 27 (331) | 26 (244) | 33 (203) |
| Monthly household income, % |  |  |  |  |
| <1500 € | 16 (292) | 13 (170) | 13 (130) | 10 (69) |
| 1500-2300 € | 29 (512) | 29 (387) | 30 (302) | 27 (182) |
| 2300-3000 € | 27 (474) | 28 (378) | 30 (299) | 28 (190) |
| >3000 € | 28 (493) | 30 (398) | 28 (282) | 34 (232) |
| Mother consuming tobacco during pregnancy, % | 26 (457) | 22 (294) | 23 (229) | 20 (135) |
| Mother consuming alcohol during first trimester, % | 43 (775) | 45 (593) | 46 (468) | 42 (282) |
| Maternal depression score during pregnancy | 11.6 (8.0) | 11.1 (7.7) | 11.0 (7.5) | 10.7 (7.3) |
| Boys | 53 (943) | 52 (701) | 54 (547) | 51 (348) |
| Gestational age, weeks | 39 (2) | 39 (2) | 39 (2) | 39 (2) |
| Any breastfeeding duration (months), % |  |  |  |  |
| Never | 28 (489) | 27 (358) | 29 (295) | 0 (1) |
| < 3.0 | 26 (470) | 27 (368) | 27 (273) | 35 (235) |
| 3.0-6.0 | 23 (403) | 21 (283) | 20 (209) | 30 (203) |
| ≥ 6.0 | 23 (415) | 25 (331) | 24 (243) | 35 (237) |
| Childcare arrangements |  |  |  |  |
| Nursery/nanny | 60 (898) | 65 (875) | 66 (676) | 68 (459) |
| Family member (mother included) | 40 (605) | 35 (466) | 34 (345) | 32 (218) |

^1^ Values are mean ± SD or % (n).

Abbreviations: CDI-2: communicative development inventory at age 2 years, BMI: body mass index
