## Supplemental Figure 4 for "Associations between perinatal biomarkers of maternal dairy fat intake and child cognitive development: results from the EDEN mother-child cohort"

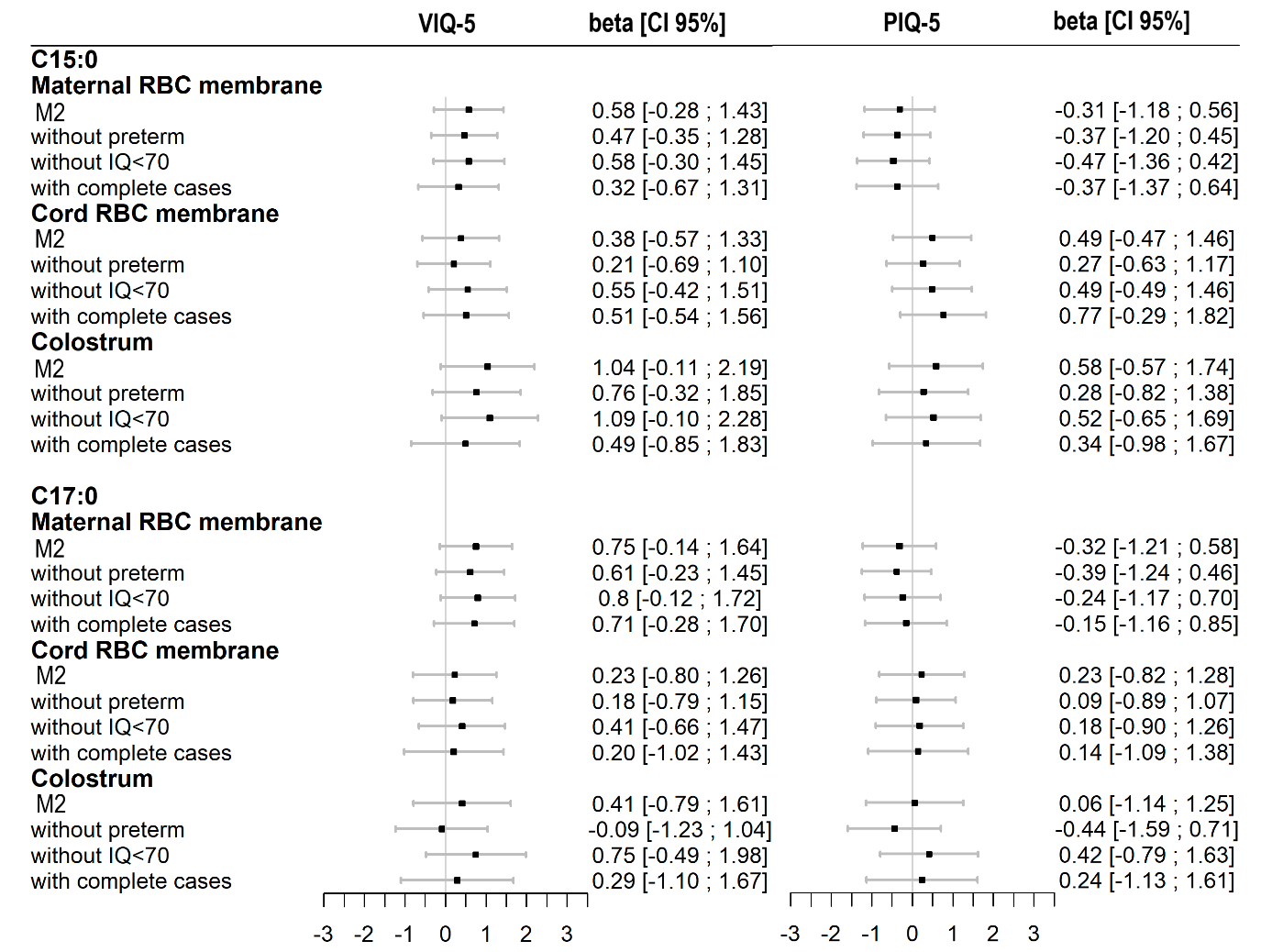

**Supplemental Figure 4**. Associations between C15:0 and C17:0 levels in perinatal biofluids and child’s VIQ-5 (on the left) and PIQ-5 at age 5-6 years (on the right) after removing children born preterm, with an IQ<70 or with missing data for covariates. For both cognitive outcomes, M2 was run on n=1082, n=821 and n=547 for maternal, cord RBC membranes and colostrum. For both cognitive outcomes, models were linear regressions. Values are β [CI95%] expressed as 1SD change in VIQ-5/ PIQ-5 per 1SD increase in C15:0 or C17:0 levels (standardized for comparison).

Models were adjusted for study center, maternal age at delivery, child’s sex, maternal pre-pregnancy BMI, parity, tobacco and alcohol consumption during pregnancy, depression score during pregnancy, monthly household income, parental education attainment, child main caregiver at age 2 years, stimulation activities level at age 5-6 years and maternal healthy diet score. Models for colostrum, were additionally adjusted for gestational age, the day of collection and any breastfeeding duration. M2 was further run after removing children born preterm (for maternal RBC, n=1,021; for cord RBC, n=785; for colostrum, n=523), children with an IQ<70 (for maternal RBC, n=1,063; for cord RBC, n=806; for colostrum, n=540) and children with missing data for covariates (for maternal RBC, n=820; for cord RBC, n=625; for colostrum, n=418).
