## Supplemental Figure 2 for "Associations between perinatal biomarkers of maternal dairy fat intake and child cognitive development: results from the EDEN mother-child cohort"

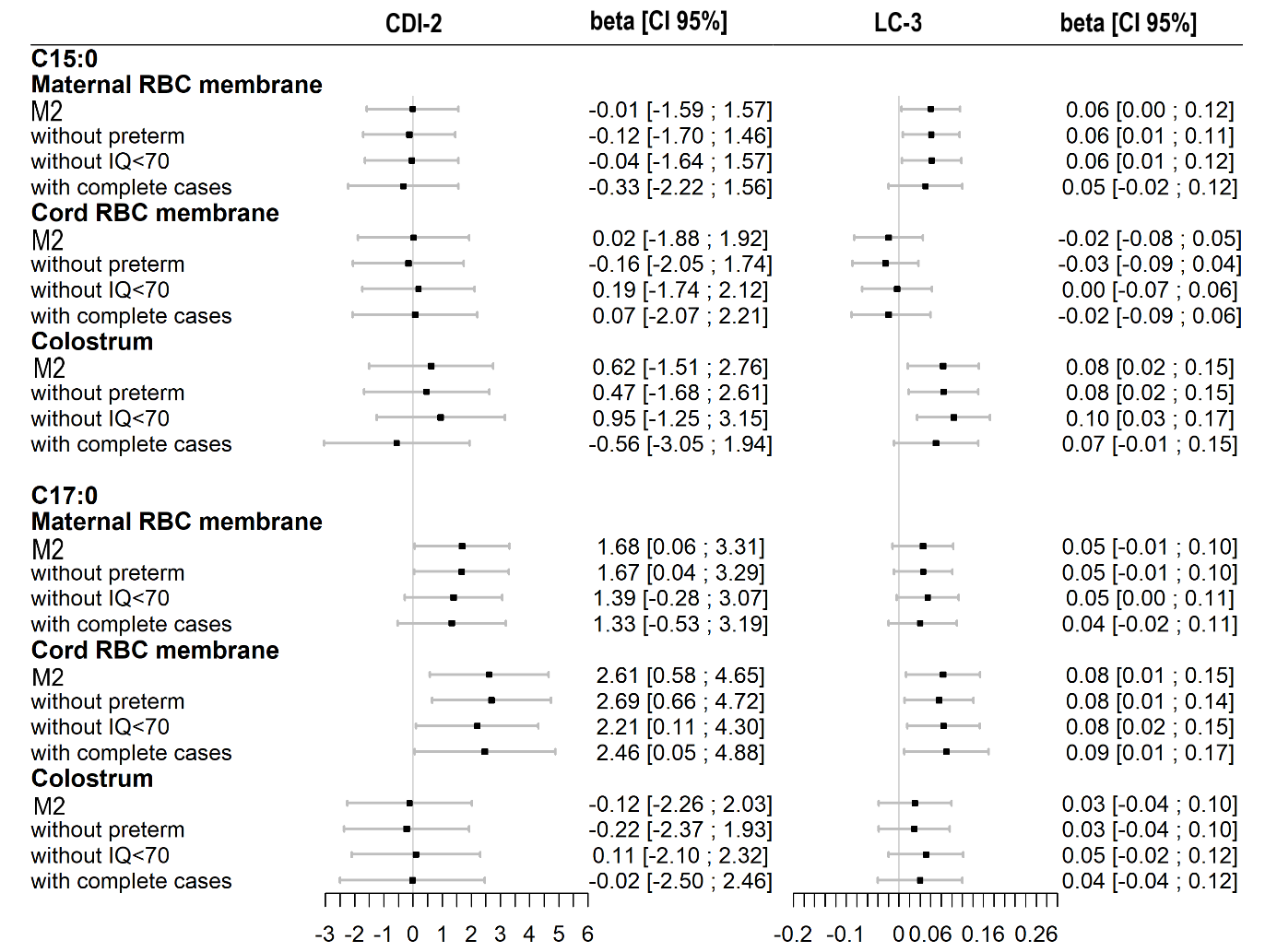

**Supplemental Figure 2.** Associations between C15:0 and C17:0 levels in perinatal biofluids and child’s language abilities at ages 2 (CDI-2; on the left) or 3 years (LC-3; on the right) after removing children born preterm, with an IQ<70 or with missing data for covariates. For CDI-2, M2 was run on n=1397, n=1060 and n=710 for maternal, cord RBC membranes and colostrum, respectively. For LC-3, M2 was run on n=1220, n=927 and n=621 for maternal, cord RBC membranes and colostrum, respectively. For both cognitive outcomes, models were linear regressions. Values are β [CI95%] expressed as 1 point (CDI-2) or 1SD (LC-3) change in cognitive outcomes per 1SD increase of C15:0 or C17:0 levels (standardized for comparison). Models (M2) were adjusted for study center, maternal age at delivery, child’s sex, age at cognitive assessment (only in models with CDI-2) maternal pre-pregnancy BMI, parity, tobacco and alcohol consumption during pregnancy, depression score during pregnancy, monthly household income, parental education attainment, child main caregiver at age 2 years, stimulation activities level at age matched with the studied outcome and preschool attendance (only in models with LC-3) and maternal healthy diet score. Models for colostrum, were additionally adjusted for gestational age, the day of collection and any breastfeeding duration. M2 was further run after removing children born preterm (for maternal RBC, n=1,320-1,155 (CDI-2-LC-3); for cord RBC, n=1,015-883; for colostrum, n=677-594), children with an IQ<70 (for maternal RBC, n=1,375-1,203; for cord RBC, n=1043-917; for colostrum, n=703-617) and children with missing data for covariates (for maternal RBC, n=1105-930; for cord RBC, n= 841-705; for colostrum, n= 565-480).
